# Evaluation of cold pain tolerance in patients with fibromyalgia and opioid use by survival analysis

**DOI:** 10.1101/2022.03.26.22272966

**Authors:** Eden Z. Deng, Daniel P. Weikel, Katherine T. Martucci

**Author notes:** **Corresponding Author:** Katherine T. Martucci, Director, Human Affect and Pain Neuroscience (HAPN) Lab, Duke University Medical Center, Box DUMC 3094, Durham, NC 27710 USA. **Funding:** This project was funded by the National Institutes of Health, National Institute of Drug Abuse (NIDA), K99/R00 DA040154 (awarded to K.T.M.) and the Department of Anesthesiology, Duke University Medical Center.

## Abstract

**Purpose:** Cold pain tolerance is traditionally measured using the cold pressor test (CPT), a clinical pain research method in which participants immerse an extremity (i.e., hand or foot) into cold water for as long as tolerable. Prior research studies have investigated cold pain tolerance in patients with chronic pain, yet few have used survival analysis, which allows for proper handling of data censoring and therefore, the optimal statistical method for CPT data analysis. The goal of the present study was to use survival analysis to evaluate cold pain tolerance in patients with fibromyalgia. Furthermore, we aimed to model relationships between psychological and clinical variables as well as opioid medication use and cold pain tolerance.

**Patients and Methods:** A total of 85 patients with fibromyalgia (42 who were taking opioids) and 47 healthy pain-free controls were involved in two studies that provided CPT and questionnaire data. Survival analysis using Cox regression models evaluated group effects (patients vs. controls) and effects of psychological, clinical, and medication factors on cold pain tolerance.

**Results:** Patients with fibromyalgia, as compared to healthy controls, exhibited significantly lower CPT survival (HR = 2.17, 95% CI: [1.42, 3.31], p = 0.00035). Cox regression models indicated that the psychological and clinical measures included did not strongly mediate this relationship (p > 0.05). When comparing across patient cohorts (Study 1 vs. Study 2), results were consistent across non-opioid-taking patient and healthy control groups, while differences in CPT survival were observed across the groups of patients taking opioid pain medications.

**Conclusion:** Reduced cold pain tolerance in patients with fibromyalgia is identified by survival analysis, an optimal method for time-to-event pain measures. While our selected psychological and clinical measures do not appear to be significantly associated with cold pain tolerance, opioid medication use may impart greater cold pain tolerance among some patients.

## 1. Introduction

The prevalence of chronic pain has increased across all demographics and, in conjugation with the current opioid epidemic, imparts a major burden on society.^1^ While the complex and interacting multi-body system processes that drive chronic pain remain unclear, pain tolerance is one aspect of characterizing chronic pain that may provide useful insights to underlying and differential mechanisms (i.e., among individual patients and between different types and stages of chronic pain). Cold pain tolerance, specifically, can be measured by the cold pressor test (CPT), which has been used in clinical settings and in pain research to measure pain tolerance and sensitivity in a broad range of healthy and patient populations.^2^

Despite many prior published studies which include CPT data, most published studies using CPT have been analyzed without proper handling of censoring in the data (i.e., with parametric statistical methods).^3^ In particular, published analyses of pain tolerance in chronic pain populations have rarely used survival analysis methods, despite this approach being more suitable for the censored data generated by pain tolerance tests. Notably, a few studies have utilized survival analysis for CPT data to study pain tolerance in adults with irritable bowel syndrome (IBS),^4^ to study psychological and genetic predictors of pain tolerance in healthy subjects,^5^ and to measure opioid-induced hyperalgesia (OIH) in opioid-dependent patients.^6^ However, regarding pain tolerance in the fibromyalgia patient population, to our knowledge, CPT results have not been reported with survival analysis methods.

Data produced by the CPT assessment are “censored time-to-event data” which are appropriately analyzed by regression models, the most common of which is the Cox proportional-hazards model. The Cox proportional-hazards model is a semiparametric model that predicts the hazard function from a set of time-independent explanatory variables.^7^ In the case of data acquired via CPT assessment, the hazard function *h(t)* represents an individual’s risk of removing their hand from the cold water at some time *t* during the CPT. The hazard ratio (HR) quantifies the difference in the hazard experienced by two groups at any given time.

Pain tolerance may be affected by numerous health and psychological factors in both healthy individuals and patients with chronic pain, such as fibromyalgia. Prior literature indicates that, compared to healthy individuals, patients with fibromyalgia demonstrate less tolerance to cold pain.^8–10^ Further, such CPT differences may be modulated by changes in activity within brain sensorimotor, attentional, and executive control networks, as well as subcortical/brainstem areas.^11–14^ In both pain-free individuals and individuals with chronic pain, state anxiety has been linked to greater pain sensitivity from a variety of noxious stimuli.^15–18^ In patients with fibromyalgia, greater clinical pain intensity relates to greater number of pain areas across the body and increased negative affective states.^19^ Moreover, there is evidence that positive affect^20,21^ or affect balance^22^ may moderate the association between negative affect and pain intensity.

Pain tolerance may be influenced by the use of opioid medications. Opioid medications prescribed as long-term treatment for chronic pain pose risks of addiction, overdose, and opioid-induced hyperalgesia (i.e., increased pain response when taking opioids).^23,24^ Concurrent with mechanisms of opioid-induced hyperalgesia, opioid use may affect pain tolerance in chronic pain patients. Indeed, opioid-dependence has been shown to correlate with greater sensitivity to cold pain,^25^ and the CPT has been used as a measure of opioid-induced hyperalgesia in patients with fibromyalgia and other opioid-dependent patients.^6,26^

The aim of the present study was to provide an analysis of CPT-evaluated cold pain tolerance in patients with fibromyalgia with enhanced rigor and reliability by involving survival analysis statistical methods and *a priori* power analysis. We hypothesized that patients with fibromyalgia would demonstrate less cold pain tolerance (i.e., higher hazard of CPT survival) compared to healthy pain-free controls. We additionally hypothesized that factors of state anxiety, negative affect, number of pain areas in the body, and opioid use would be related to less pain tolerance in patients with fibromyalgia. Lastly, we explored additional psychological and clinical factors in their relationship to cold pain tolerance in patients with fibromyalgia.

## 2. Methods

### 2.1. Participants

All patient and control participants were female and participated in one of two separate studies during which data were collected. The two independent data collection sources were conducted by different research teams at different times and locations. CPT data from 35 patients and 17 controls, recruited from the regions surrounding Palo Alto, CA, were collected between 2015 and 2018 for Study 1 which was conducted at Stanford University. Results from analyses of other non-CPT-related data collected from Study 1 have been reported previously.^27–29^ Data from 50 patients and 30 controls, recruited from the regions surrounding Durham, NC, were collected between 2019 and 2021 for Study 2 which was conducted at Duke University.

In Study 1, all fibromyalgia patients met the modified American College of Rheumatology (ACR) 2011 criteria for fibromyalgia [widespread pain index (WPI) ≥ 7 + symptom severity (SS) ≥ 5, or WPI 3–6 + SS ≥ 9; symptoms present at a similar level for at least 3 months; no disorder that would otherwise explain the pain].^30^ In Study 2, fibromyalgia patients met the revised ACR 2016 criteria which includes a slight change in WPI criteria (range 4–6).^31^ All fibromyalgia patients were required to have pain in all four quadrants of the body, have an average pain score over the past month of at least 2 on a 0-10 verbal scale, have no MRI contraindications, and not be pregnant or nursing. Healthy individuals were required to have no history of chronic pain, not be pregnant or nursing, have no MRI contraindications, not be taking pain- or mood-altering medications at the time of the study, and have no depression or anxiety disorder. In total, 85 fibromyalgia patients and 47 healthy participants signed written informed consent indicating their willingness to participate in the study, understanding of all study procedures, and acknowledgement that they could withdraw from the study at any time. All procedures conducted were in accordance with the Declaration of Helsinki and were approved by the Stanford University Institutional Review Board for Study 1 and the Duke University Institutional Review Board for Study 2. A data use agreement was established allowing the data collected at Stanford to be analyzed by the research team at Duke University (K.T.M. was the PI for both Study 1 and Study 2). Pre-registered data analysis plans including hypotheses and exploratory analyses were published on the Open Science Framework (OSF) website at: DOI: 10.17605/OSF.IO/H2KGW (https://archive.org/details/osf-registrations-h2kgw-v1).

### 2.2. Medication usage

A total of 42 patients with fibromyalgia (18 from Study 1; 24 from Study 2) were enrolled based on their use of prescribed opioid medications as part of their ongoing pain treatment. These 42 participants were required to have been taking opioid medications for at least 3 months prior to and at the time of their participation in the study. The remaining 43 fibromyalgia participants had never taken opioids for a period greater than 30 days, had not taken any opioids within the 90 days before study participation, and were not taking opioid medications as part of their pain treatment at the time of their study participation. Opioid use and additional information about medication dosage and duration of use were recorded, and all participants were allowed to continue their medication use as normal during their participation in the study (see **Table 2** and **Table 5**).

### 2.3. Study procedures

#### 2.3.1. Sample size

The total required sample size of the combined (Study 1 + Study 2) dataset was determined based on the data collected in Study 1. Using the powerSurvEpi package in R, a sample size for Cox regression was calculated based on the hazard ratio (HR) between the fibromyalgia group and the healthy control group in the CPT data from Study 1 (HR = 2.83). To obtain a minimum power of 0.80 at the 0.05 alpha level for a targeted HR of 2.0 in the combined (Study 1 + Study 2) dataset, it was determined that the combined dataset should have a minimum healthy control sample size of 39 (expecting 28 uncensored events; i.e., time < 120 seconds) and fibromyalgia sample size of 79 (expecting 73 uncensored events). Thus, the final combined study sample used in the analysis contained sufficient counts of healthy control (n = 47) and fibromyalgia (n = 85) participants.

#### 2.3.2. Cold pressor test

The cold pressor test was conducted in different facilities and by different experimenters between Study 1 and Study 2, with slight variations in protocol. In Study 1, the initial temperature of the cold water was customized for each participant by determining and using the temperature associated with each participant’s pain intensity rating of 3 on a 0-10 visual analog scale (VAS, anchors of “no pain” and “worst pain imaginable”).^32^ This individualized temperature selection was determined prior to the CPT assessment by conducting a cold water immersion test of the left hand, up to 30 seconds in duration and repeated up to 2 times as necessary at different temperatures with 5 minutes of rest between repeated tests. In Study 2, the temperature of the cold water was standardized among all participants to 5º C. For both studies, the CPT assessment was conducted by first filling a plastic container with ice water, then adjusting and recording the temperature of the water. Participants were asked to submerge their right hand in the cold water (with all ice removed) for as long as tolerable. The experimenters started a timer as soon as the participant first immersed her hand into the water and allowed the participant to keep her hand in for up to 2 minutes. After the cold pressor test, each participant was asked to rate her pain intensity and unpleasantness associated with the cold water experience on a 0-10 VAS sliding scale.

#### 2.3.3. Psychological and clinical questionnaires

In addition to providing demographic and medication information, all subjects completed the following set of questionnaires: Beck Depression Inventory (BDI),^33^ Behavioral Inhibition System/Behavioral Approach System (BIS/BAS),^34^ Positive and Negative Affect Schedule (PANAS),^35^ Profile of Mood States (POMS),^36^ State-Trait Anxiety Inventory (STAI-State, STAI-Trait),^37^ Brief Pain Inventory (BPI),^38^ Fibromyalgia Assessment Status (FAS),^39^ and Patient-Reported Outcome Measurement Information System (PROMIS) Fatigue.^40^

### 2.4. Statistical analyses

Questionnaire data were stored in and downloaded from secure REDCap databases. CPT data that were documented on case report forms were entered manually (by double data confirmatory entry) into Excel spreadsheets prior to analysis. All analyses were pre-registered in the OSF registry (DOI: 10.17605/OSF.IO/H2KGW) and were conducted in R version 3.6.0.

Because the CPT produces time-to-event data with right-censoring, the data were fit with Cox proportional-hazards models predicting participants’ hazard in the CPT from their group status (patient or control), demographic information, behavioral and psychological measures, and medication information. Prior to analysis, variables were assessed for greater than 5% missingness to determine whether data imputation would be required. Missing values were not imputed for variables with less than 5% missingness. All models used in this analysis contained an adjustment variable for starting temperature and a binary variable indicating Study (Study 1 or Study 2). Nested Cox models were compared using the likelihood ratio test (LRT), and non-nested Cox models were compared using the plrtest function in R (created by Thomas Hielscher) which implements a partial LRT suited for non-nested Cox models.^41^

Cold pain tolerance was first compared between patient (i.e., combined opioid and non-opioid patients) and healthy control groups using a Cox model predicting CPT hazard from fibromyalgia group status. To investigate the effect of demographic and psychological/clinical measures on the relationship between fibromyalgia and CPT hazard, a multivariate Cox model was built by selecting up to 4 additional covariates to add to the model in addition to group status (**Figure 1A**). The 4 additional covariates were first selected based on preliminary significance with CPT hazard (described below) from an array of *a priori* identified possible covariates, including age, race (White, Asian, Black, other), BDI score, BAS reward responsiveness, BAS drive, BAS fun-seeking, BIS score, PANAS positive affect, PANAS negative affect, POMS total mood disturbance, STAI state anxiety, and STAI trait anxiety. The CPT hazard function given by each covariate was first modeled individually (including an adjustment variable for starting temperature and a binary variable for Study), and then up to 4 of the most significant variables with coefficient p < 0.2 were selected. These variables were tested for high correlation (continuous variables: Pearson r > 0.7, categorical variables: *χ*^2^ test p < 0.01), and only the most significant variable of any group of highly correlated variables was included in the model.

**Figure 1:**
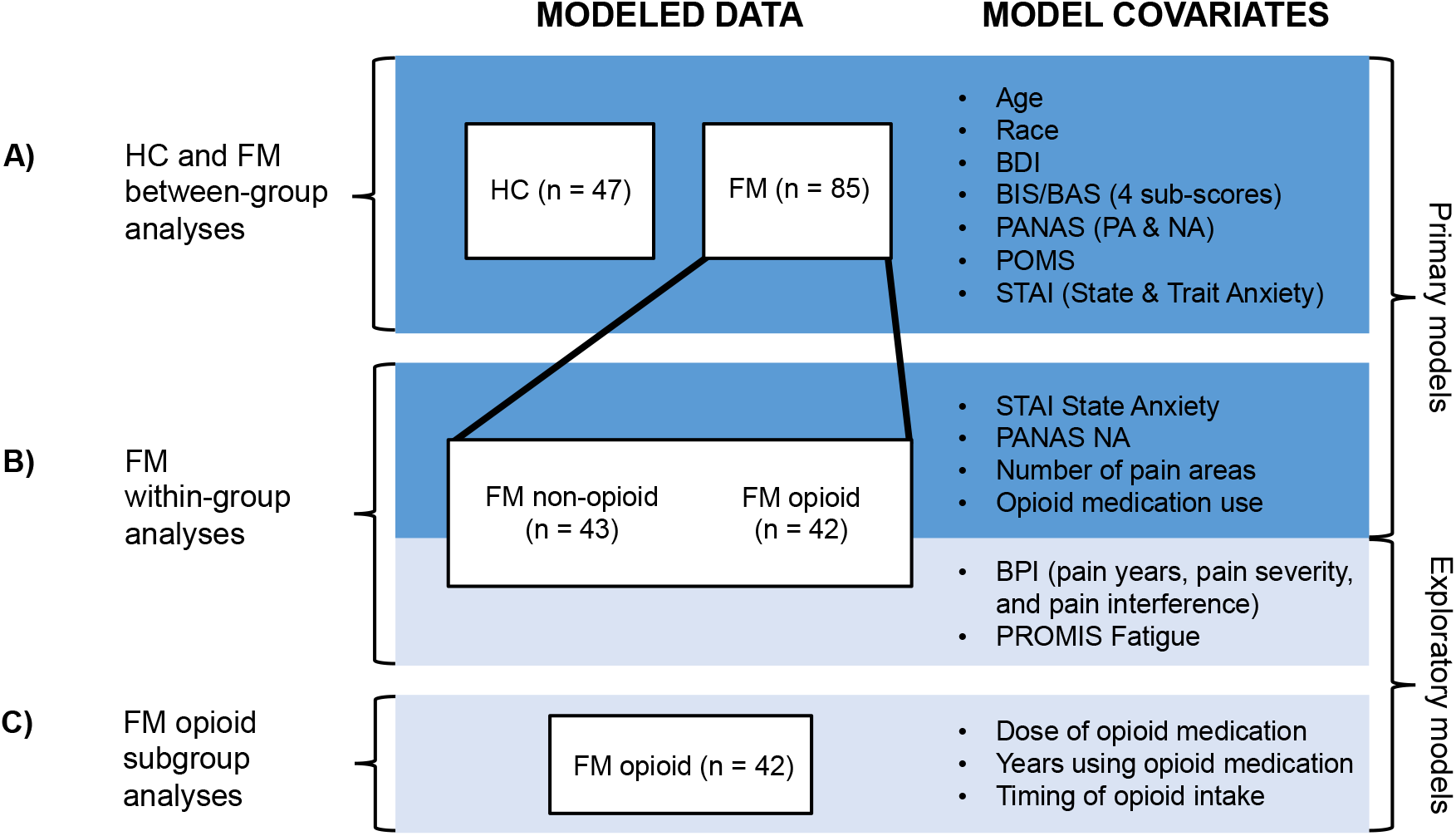
Diagram of statistical models. The primary hypotheses were first tested (A) across all participants (both healthy control [HC] and fibromyalgia [FM] groups) and (B) within the fibromyalgia group. Exploratory tests were then conducted (B) within the fibromyalgia group as well as (C) within the opioid-taking subgroup. All tests were conducted using Cox proportional hazards models. Abbreviations: BDI, Beck Depression Inventory; BIS/BAS, Behavioral Inhibition System/Behavioral Activation System; PANAS, Positive and Negative Affect Schedule; POMS, Profile of Mood States; STAI, State-Trait Anxiety Inventory; BPI, Brief Pain Inventory; PROMIS, Patient-Reported Outcome Measurement Information System.

State anxiety, negative affect, number of pain areas in the body, and opioid medication use were hypothesized to decrease pain tolerance in patients with fibromyalgia. To test these hypotheses within the patient group, the CPT hazard function was modeled by each including STAI state anxiety, PANAS negative affect, FAS pain areas, and a binary variable indicating opioid medication use (**Figure 1B**).

Due to non-normality of the data, all group comparisons of the psychological/clinical variables were conducted using the Mann-Whitney Wilcoxon Test. The cox.zph function in R was used to confirm proportional hazards for all models. Observations with deviance residuals greater than 2.5 or less than -2.5 were identified as possible outliers.

### 2.5. Exploratory analyses

To validate the multivariate model and address the possibility of correlated groups of covariates in the model, Lasso regression was also used to identify variables predictive of CPT hazard. The Lasso approach tends to select one predictor out of a group of correlated predictors and discard the rest, and it can be more accurate than stepwise selection.^42^

In addition to state anxiety, negative affect, number of body areas with pain, and opioid medication use, other factors potentially predictive of CPT survival within the patient group were also explored with Cox models. These exploratory variables included the covariates tested in the primary analysis as well as patients’ number of years with pain (BPI), average pain intensity (BPI), average pain interference (BPI), and PROMIS Fatigue score (**Figure 1B**). Because the study effect on CPT survival was significant, Study 1 and Study 2 were also separately analyzed for these relationships. Additionally, a stronger relationship between opioid medication use and CPT survival was observed in Study 2 compared to Study 1; to determine whether opioid dosage, duration of use, or timing of last intake (the opioid half-life range during which the CPT test was performed, calculated based on the time of last opioid dose and the drug-specific half-life) might account for this between-study difference observed in the opioid-taking population, a sub-analysis of the opioid-taking fibromyalgia group was conducted by testing for study differences in these three measures and separately testing them as predictors of CPT survival (**Figure 1C**). Lastly, a *post-hoc* sensitivity analysis of opioid medication effects was conducted by re-evaluating the primary results on the dataset including only non-opioid-taking patients and healthy controls.

## 3. Results

### 3.1. Participants

Data from a total of 85 patients and 47 controls (see **Table 1** for demographic information) were analyzed. Forty-three out of the 85 fibromyalgia participants were not taking opioid medications and were taking over-the-counter pain medications. The remaining 49% (n = 42) of the fibromyalgia patients were taking opioid pain medications. This sub-group had a median daily morphine equivalent dose (MED) of 20 mg morphine and a median opioid-taking duration of 5 years. Of the participants with fibromyalgia, 59% (n = 50) were taking anti-depressants, 26% (n = 22) were taking benzodiazepines or benzodiazepine-like medications, and 56% (n = 48) were taking muscle relaxants or anti-seizure medications (**Table 2**).

**Table 1:** Participant demographics. One control participant did not indicate handedness. The race category labeled “other” includes participants of Hispanic or Latinx ethnicity. No significant age difference was observed between the patient group and control group (Mann-Whitney p = 0.84).

|  |  | **Patients** | **Controls** |
| --- | --- | --- | --- |
| **Total Participants** |  | 85 | 47 |
| **Right Handed** |  | 79 | 44 |
| **Self-Identified Race** | *Asian* | 3 | 10 |
|  | *White/Caucasian* | 67 | 33 |
|  | *Black/African American* | 9 | 2 |
|  | *Other* | 6 | 2 |
| **Hispanic or Latinx Ethnicity** |  | 9 | 2 |
| **Median Age** |  | 47 | 45 |

**Table 2:** General medications. The number of fibromyalgia patients and healthy control participants (from both Study 1 and Study 2) taking each medication type is shown. One control participant with premenstrual symptoms (2 days per month) reported taking gabapentin (100 mg/day, 2 days per month) and fluoxetine (40 mg/day), one control participant reported taking naproxen for menstrual cramps as needed, and another control participant reported taking celecoxib (200 mg) 3 weeks prior to the study visit due to a sports-related ankle injury. One control participant reported taking zolpidem for sleep, and another control participant reported taking cannabidiol oil (0.5 mg/day) for better rest. Abbreviations: Nonsteroidal anti-inflammatory drug, NSAID; gamma-aminobutyric acid, GABA; serotonin and noradrenergic reuptake inhibitor, SNRI; selective serotonin reuptake inhibitor, SSRI; serotonin antagonist and reuptake inhibitor, SARI; norepinephrine-dopamine reuptake inhibitor, NDRI.

|  | **Patients** | **Controls** |
| --- | --- | --- |
| **Opioids** | 42 | 0 |
| *Hydrocodone/acetaminophen* | 16 |  |
| *Tramadol* | 15 |  |
| *Oxycodone/acetaminophen* | 4 |  |
| *Morphine* | 3 |  |
| *Codeine/acetaminophen* | 2 |  |
| *Tapentadol* | 2 |  |
| *Hydrocodone* | 1 |  |
| *Oxycodone* | 1 |  |
| *Methadone* | 1 |  |
| *Hydromorphone* | 1 |  |
| **NSAID** | 37 | 2 |
| **Acetaminophen** | 30 | 0 |
| **GABA Analogue** (e.g., gabapentin) | 28 | 1 |
| **Muscle Relaxant** (e.g., cyclobenzaprine) | 27 | 0 |
| **SNRI** (e.g., duloxetine) | 26 | 0 |
| **Benzodiazepine** | 16 | 0 |
| **Antiepileptic** (e.g., topiramate) | 15 | 0 |
| **SSRI** (e.g., fluoxetine) | 14 | 2 |
| **Triptans** (e.g., sumatriptan) | 13 | 0 |
| **SARI** (e.g., trazodone) | 10 | 0 |
| **NDRI** (e.g., methylphenidate, bupropion) | 10 | 0 |
| **Ondansetron** | 7 | 0 |
| **Other Anxiolytic** (e.g., buspirone) | 6 | 0 |
| **Benzodiazepine-like** (e.g., eszopiclone, zolpidem) | 5 | 1 |
| **Medical Cannabis** | 5 | 1 |
| **Tricyclic Antidepressant** (e.g., amitriptyline) | 5 | 0 |
| **Antipsychotic** (e.g., lurasidone, prochlorperazine) | 4 | 0 |
| **Low Dose Naloxone** | 3 | 0 |
| **Topical Lidocaine Patch** | 1 | 0 |

### 3.2. Psychological and clinical measures

Less than 5% of the questionnaire responses were missing from the data. Among the patient group, participants’ reported number of body areas (FAS) with pain ranged from 3 (the minimum criteria) to 19 (maximum). The median pain severity was 5.8/10 and median pain interference was 6.4/10 (BPI). The patient vs. healthy control groups showed significant differences in CPT time, STAI trait anxiety, STAI state anxiety, PANAS positive affect, PANAS negative affect, BAS fun-seeking, BIS score, POMS total mood disturbance, and BDI score (**Table 3**).

**Table 3:** Clinical and psychological measures between groups. Participant counts for each measure differ from the total number of participants (healthy controls [HC] n = 47, fibromyalgia [FM] n = 85) because some participants did not complete all questionnaires. Abbreviations: BDI, Beck Depression Inventory; BAS, Behavioral Activation System; BIS, Behavioral Inhibition System; PANAS, Positive and Negative Affect Schedule; POMS, Profile of Mood States; STAI, State-Trait Anxiety Inventory; BPI, Brief Pain Inventory; FAS, Fibromyalgia Assessment Status; PROMIS, Patient-Reported Outcome Measurement Information System; IQR, interquartile range. Data are presented for descriptive purposes only, therefore the Mann-Whitney p-values shown are not corrected for multiple comparisons.

|  | Patients |  |  | Controls |  |  | Mann-Whitney<br>p-value |
| --- | --- | --- | --- | --- | --- | --- | --- |
|  | N | Median | IQR | N | Median | IQR |  |
| CPT Time | 85 | 23.12 | [12.00, 41.00] | 47 | 38.62 | [23.425, 120.00] | < 0.001 |
| BDI Depression | 84 | 16.00 | [11.00, 24.00] | 47 | 1.00 | [0.00, 3.00] | < 0.001 |
| BAS Reward Responsiveness | 81 | 17.00 | [15.00, 18.40] | 46 | 17.00 | [15.40, 18.00] | 0.707 |
| BAS Drive | 81 | 11.00 | [9.00, 13.00] | 46 | 11.10 | [9.25, 13.00] | 0.356 |
| BAS Fun-Seeking | 81 | 10.40 | [9.00, 12.00] | 46 | 12.00 | [10.55, 13.00] | 0.003 |
| BIS Behavioral Inhibition | 81 | 22.00 | [19.00, 24.00] | 46 | 18.70 | [16.20, 21.60] | < 0.001 |
| PANAS Positive Affect | 85 | 25.00 | [19.00, 31.00] | 47 | 35.00 | [32.00, 40.50] | < 0.001 |
| PANAS Negative Affect | 85 | 20.00 | [16.00, 26.00] | 47 | 12.00 | [11.00, 15.00] | < 0.001 |
| POMS Total Mood Disturbance | 85 | 18.00 | [9.00, 36.00] | 47 | -4.00 | [-8.00, 2.00] | < 0.001 |
| STAI State Anxiety | 85 | 41.00 | [35.00, 47.00] | 46 | 26.00 | [22.00, 32.00] | < 0.001 |
| STAI Trait Anxiety | 81 | 45.00 | [40.00, 54.00] | 47 | 31.00 | [25.50, 36.00] | < 0.001 |
| BPI Pain Years | 82 | 10.00 | [5.00, 15.00] |  |  |  |  |
| BPI Pain Severity | 85 | 5.75 | [4.75, 6.50] |  |  |  |  |
| BPI Pain Interference | 83 | 6.43 | [5.214, 7.50] |  |  |  |  |
| FAS Total Pain Areas | 85 | 13.00 | [9.00, 16.00] |  |  |  |  |
| PROMIS Fatigue | 84 | 66.05 | [63.90, 72.32] |  |  |  |  |

### 3.3. Group comparison of CPT survival

The distributions of CPT time by group are shown in **Figure 2** (Mann-Whitney p = 0.00080). Fibromyalgia patients had significantly lower CPT survival probability compared to controls (HR = 2.17, 95% CI: [1.42, 3.31], p = 0.00035), when including adjustment variables for Study 1 vs. Study 2 and CPT starting temperature (**Figure 3, Table 6**).

**Figure 2:**
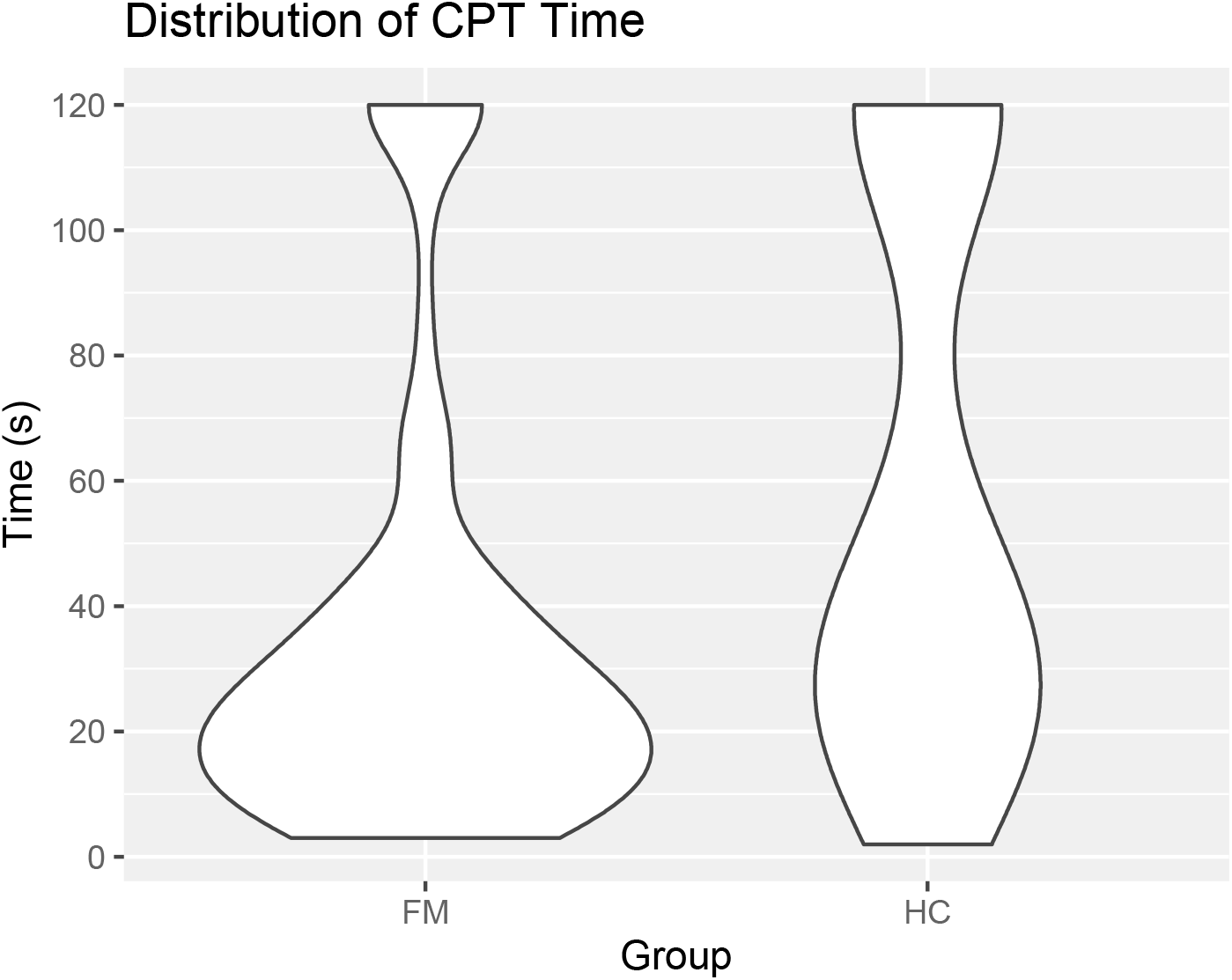
Distribution of CPT time by group. Distributions of CPT times from the fibromyalgia (FM) and healthy control (HC) groups are shown. The control group contained a larger proportion of censored times (CPT cut-off at 120 seconds) compared to the fibromyalgia group.

**Figure 3:**
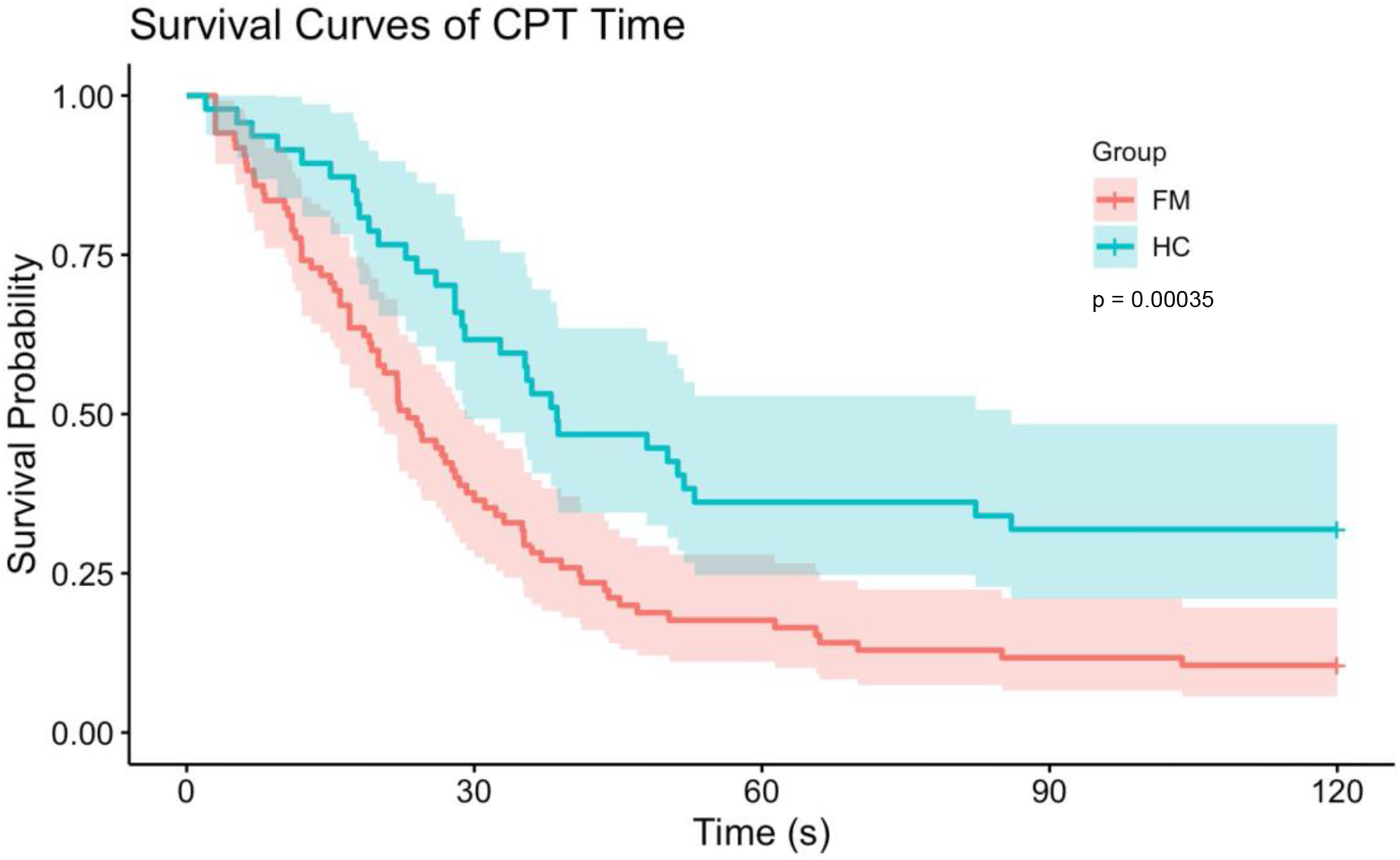
Kaplan-Meier curves of CPT assessment results by group. Kaplan-Meier curves showing CPT survival with 95% confidence intervals are shown for the fibromyalgia (FM) group and healthy control (HC) group. The p-value shown is for the group effect in the Cox regression model of CPT survival time with adjustments for starting temperature and study effect. Time-to-event data was censored at 120 seconds, and proportional hazards criteria were met for Cox regression.

Regarding study-specific effects, between the fibromyalgia participants in Study 1 and Study 2, there was a significant difference in CPT time (p = 0.00079). The patient group of Study 2 was characterized by a higher proportion of censored CPT times (16%) compared to Study 1 (3%). The healthy control group, however, showed no significant differences in CPT time between studies.

Results from the single-variable Cox models of CPT hazard, which each included a psychological/clinical variable, showed that none were significantly associated with CPT survival (p > 0.05) after accounting for starting temperature, study differences, and group status. However, both Study 1 and fibromyalgia group status were significantly associated with greater hazard in each of these models. The multivariate Cox model included the variable for patient group status (HR = 1.99, p = 0.0020), selected variables which included BAS fun-seeking (HR = 0.94, p = 0.28), BAS drive (HR = 0.97, p = 0.50), and “other” race (HR = 1.78, p = 0.15), as well as an indicator variable for Study 2 and an adjustment variable for starting temperature (exact, measured before the CPT assessment). The multivariate model did not improve the fit for the CPT data compared to the simpler model with no psychological predictors (LRT: p = 0.16).

Variable selection with Lasso regression revealed similar results: the binary variables for patient group status (HR = 1.71, p = 0.048) and Study 2 (HR = 0.61, p = 0.017) were significant covariates selected by Lasso (10-fold cross-validation using Harrell’s C-index, and coefficients were extracted at the value of λgiving the minimum mean cross-validation error). BAS fun-seeking, BIS score, “other” race, and STAI state anxiety were also selected but were not significant predictors of CPT survival. The Lasso-selected Cox model was neither a significant improvement upon the p-value-selected multivariate Cox model (partial LRT: p = 0.82) nor upon the original simple Cox model with no psychological/clinical predictors (LRT: p = 0.23).

### 3.4. Psychological and clinical predictors of CPT survival in fibromyalgia patients

Within the patient group, STAI state anxiety, PANAS negative affect, number of pain areas, and opioid medication use were all not significantly associated with CPT survival probability (p > 0.0125). Other exploratory variables, including BAS fun-seeking (HR = 0.86, p = 0.014) and BAS drive (HR = 0.89, p = 0.021), had stronger relationships (i.e., larger effect sizes) with CPT survival in fibromyalgia patients (**Table 4**) (but not in healthy controls). The CPT survival rate in Study 2 patients was greater compared to Study 1 patients (HR = 0.48, 95% CI: [0.30, 0.77], p = 0.0026) even when accounting for CPT starting temperature. (This study difference in CPT survival was absent in the healthy control group [HR = 0.83, p = 0.62]). No psychological or clinical variables were significant predictors of CPT survival in the patient group of either study after correction for multiple comparisons. However, use of opioid medication was associated with greater CPT survival in Study 2 (HR = 0.51, 95% CI: [0.27, 0.97], p = 0.041) while its effects were unclear in Study 1 (HR = 0.88, p = 0.71), although no interaction effect was detected between study and opioid medication use (p = 0.26).

**Table 4:** Fibromyalgia group: Cox regression HRs and p-values stratified by study. Cox regression statistics within the fibromyalgia patient group (n = 85) are reported for covariates selected for *a priori* tests (opioid medication use, PANAS negative affect, number of pain areas, and STAI state anxiety) as well as exploratory covariates (BAS fun-seeking and BAS drive). Models differ in number of data points (± 4) due to participants with incomplete questionnaire responses. Because significant CPT survival differences were observed between the patient groups of Study 1 and Study 2, the effects of each covariate are also reported within each study. Each model with combined data included an adjustment variable for starting temperature and an indicator variable for Study 2. Each model with only Study 1 (n = 35) or only Study 2 (n = 50) data included an adjustment variable for starting temperature (exact, measured before CPT assessment). P-values are reported for descriptive purposes and are not corrected for multiple comparisons. Use of opioid medication was associated with greater CPT survival in Study 2 while effects were not detected in Study 1, and all other covariates tested showed similar HRs across both studies.

| Variable |  | Combined Study 1 + Study 2 |  |  | Study 1 |  |  | Study 2 |  |  |
| --- | --- | --- | --- | --- | --- | --- | --- | --- | --- | --- |
|  |  | HR | 95% CI | P-value | HR | 95% CI | P-value | HR | 95% CI | P-value |
| *A priori* | Opioid Medication Use | 0.684 | [0.433, 1.081] | 0.104 | 0.877 | [0.444, 1.731] | 0.705 | 0.509 | [0.265, 0.974] | 0.041 |
|  | PANAS Negative Affect | 0.982 | [0.952, 1.014] | 0.266 | 0.979 | [0.934, 1.027] | 0.382 | 0.987 | [0.947, 1.029] | 0.537 |
|  | FAS Number of Pain Areas | 1.033 | [0.974, 1.096] | 0.275 | 1.072 | [0.97, 1.185] | 0.173 | 1.011 | [0.941, 1.085] | 0.771 |
|  | STAI State Anxiety | 1.003 | [0.983, 1.024] | 0.742 | 1.011 | [0.972, 1.051] | 0.593 | 1.001 | [0.977, 1.025] | 0.937 |
| *Exploratory* | BAS Fun-Seeking | 0.856 | [0.757, 0.969] | 0.014 | 0.856 | [0.697, 1.051] | 0.137 | 0.867 | [0.744, 1.009] | 0.066 |
|  | BAS Drive | 0.888 | [0.803, 0.983] | 0.021 | 0.907 | [0.768, 1.073] | 0.255 | 0.878 | [0.773, 0.998] | 0.047 |

### 3.5. Between-study variability: opioid-taking fibromyalgia patient subgroup

As noted above, the relationship between opioid medication use and CPT survival among fibromyalgia patients was stronger in Study 2 compared to Study 1 (**Table 4**). In the subgroup analysis of fibromyalgia patients who were taking opioids, patients had significantly different CPT times in Study 1 vs. Study 2 (p = 0.0028). In contrast, CPT times across studies did not differ significantly for the subgroup of patients with fibromyalgia who were not taking opioids (p = 0.11) nor for the healthy control group (p = 0.52) (**Table 5**).

**Table 5:** Comparison of CPT time and opioid dose, duration, and timing across studies. Descriptive statistics and Mann-Whitney p-values for CPT times of the healthy control group (HC), non-opioid-taking fibromyalgia group (FM) and opioid-taking fibromyalgia group of each study are shown. For fibromyalgia patients taking opioid medications, the MEDs (Mann-Whitney test), duration of opioid use (Mann-Whitney test), and intake timing (i.e., opioid half-life) at the time of the CPT assessment (*χ*^2^ test) are also compared between Study 1 vs. Study 2. Two patients from Study 2 did not report their opioid medication dosage, 3 patients from Study 2 did not report their duration of opioid medication use, and 1 patient from Study 2 did not report the time of their last opioid intake. The opioid-taking patient subgroup showed significant differences in CPT time between studies, whereas the non-opioid-taking patients and healthy controls did not show significant between-study differences. Abbreviations: IQR, interquartile range; MED, morphine equivalent dose.

|  |  | Study 1 | Study 2 | P-value |
| --- | --- | --- | --- | --- |
| **CPT Time** | FM non-opioid N | 17 | 26 | 0.115 |
|  | FM non-opioid median | 13.00 | 25.58 |  |
|  | FM non-opioid IQR | [11.00, 28.00] | [13.13, 39.89] |  |
|  | FM opioid N | 18 | 24 | 0.003 |
|  | FM opioid median | 17.00 | 33.71 |  |
|  | FM opioid IQR | [9.50, 25.50] | [22.03, 107.87] |  |
|  | HC N | 17 | 30 | 0.521 |
|  | HC median | 29.00 | 44.45 |  |
|  | HC IQR | [20.00, 120.00] | [28.16, 120.00] |  |
| **Opioid Daily MED (mg)** | N | 18 | 22 | 0.956 |
|  | Median | 17.50 | 20 |  |
|  | IQR | [7.88, 39.38] | [5.00, 30.00] |  |
|  | Minimum | 1.00 | 5.00 |  |
|  | Maximum | 75.00 | 90.00 |  |
| **Opioid Duration (years)** | N | 18 | 21 | 0.966 |
|  | Median | 4.25 | 5.00 |  |
|  | IQR | [0.88, 10.00] | [3.00, 7.00] |  |
|  | Minimum | 0.25 | 0.08 |  |
|  | Maximum | 40.00 | 15.00 |  |
| **Opioid Timing (N)** | < 1 half-life | 4 | 5 |  |
|  | 1 - 2 half-lives | 6 | 1 |  |
|  | 2 - 3 half-lives | 2 | 5 |  |
|  | > 3 half-lives | 6 | 12 |  |

Although CPT times in opioid-taking fibromyalgia patients differed between Study 1 and Study 2, neither duration of opioid medication use (p = 0.97), nor daily opioid dose (p = 0.96), nor distribution of intake timing (*χ*^2^ test p = 0.091) significantly differed between studies (**Table 5**). Furthermore, none of these measures were significant predictors of CPT survival in the opioid-taking patient subgroup.

### 3.6. Post-hoc sensitivity analysis of opioid medication effects

Due to the additional between-study variation present in the subgroup of patients with fibromyalgia who were taking opioids compared to the rest of the sample (**Figure 4**), the opioid-taking subgroup was removed from the analysis to identify any changes in the group and study effects on CPT survival. After removing the opioid-taking subgroup, the Cox model indicated that CPT times were no longer different between studies (p = 0.10), the study effect was no longer significant (HR = 0.67, p = 0.11), and the group effect increased (HR = 2.70, p = 0.000054) (**Table 6**). When assessing study effects on CPT survival within each group/subgroup separately, Study 2 was associated with higher CPT survival probability in opioid-taking patients (HR = 0.40, p = 0.013), but the study effect was not significant in non-opioid-taking patients (HR = 0.60, p = 0.12) or healthy controls (HR = 0.83, p = 0.62).

**Table 6:** Group effect HRs from sensitivity analysis. The Cox regression statistics for group effect are reported for all fibromyalgia patients (FM; n = 85) vs. healthy controls (HC; n = 47) (top row), as shown in **Figure 3**, for comparison to non-opioid-taking fibromyalgia patients (n = 43) vs. healthy controls (bottom row). Due to significant differences in CPT survival between the patient groups of Study 1 and Study 2, these comparisons are also reported within each study. Each model with combined data included an adjustment variable for starting temperature and an indicator variable for Study 2. Each model with only Study 1 (n = 52) or only Study 2 (n = 80) data included an adjustment variable for starting temperature (exact, measured before CPT assessment). P-values are reported for descriptive purposes and are not corrected for multiple comparisons. The group effect was stronger when comparing the non-opioid-taking patient subgroup with healthy controls, specifically in Study 2.

| Comparison | Combined Study 1 + Study 2 |  |  | Study 1 |  |  | Study 2 |  |  |
| --- | --- | --- | --- | --- | --- | --- | --- | --- | --- |
|  | HR | 95% CI | P-value | HR | 95% CI | P-value | HR | 95% CI | P-value |
| All FM vs. HC | 2.169 | [1.419, 3.315] | 0.000348 | 3.020 | [1.491, 6.116] | 0.00215 | 1.803 | [1.037, 3.136] | 0.0368 |
| FM non-opioid vs. HC | 2.695 | [1.666, 4.361] | p < 0.0001 | 3.404 | [1.522, 7.613] | 0.00286 | 2.375 | [1.291, 4.370] | 0.00541 |

**Figure 4:**
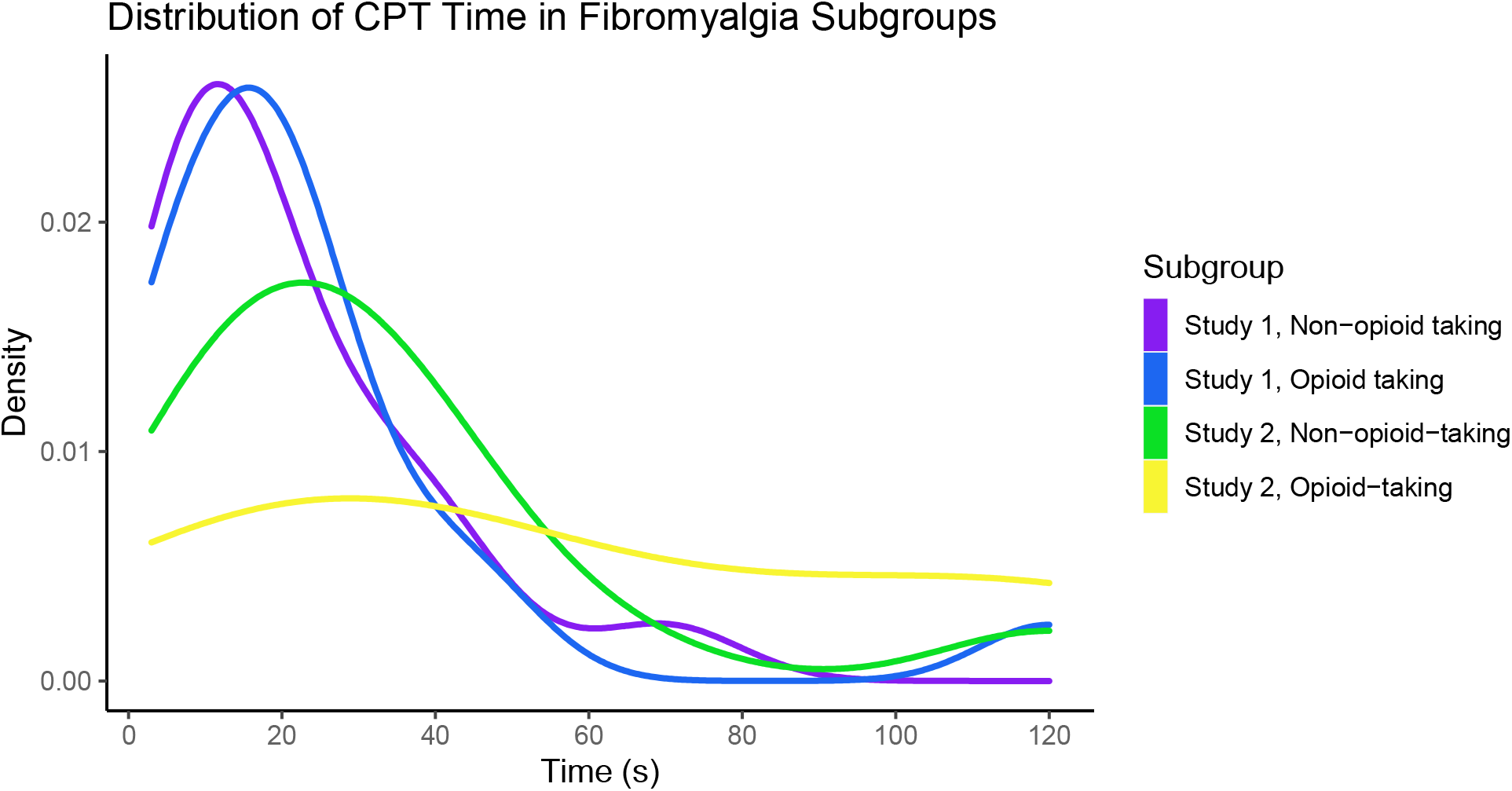
Distributions of patient CPT times by study and opioid medication use. CPT times of fibromyalgia patients in Study 1 and Study 2 are shown by subgroup (opioid-taking vs. non-opioid-taking). The distribution of CPT times for opioid-taking patients in Study 2 is shown to be more uniform compared to the right-skewed distributions of CPT times in all other fibromyalgia subgroups.

## 4. Discussion

To our knowledge, we present the first survival analysis and Cox regression modeling of CPT data in patients with fibromyalgia vs. healthy controls across combined datasets, and evaluation of cold pain tolerance differences in patients with fibromyalgia who take opioids vs. those who do not take opioids. This analysis was conducted to respond to the need to re-evaluate the statistical methods used in human and clinical pain tolerance research. Most previous analyses of CPT data have used parametric methods to form conclusions from the data. However, such approaches do not account for censored data (e.g., CPT time), which are almost always not normally distributed and cannot be effectively normalized with data transformations.^3^ Furthermore, multivariate methods (as opposed to single-variable comparisons) are needed to more accurately model the complex, interacting processes involved in pain tolerance. Here, we analyzed CPT data with the Cox proportional-hazards model, a semiparametric model widely used in various disciplines to study right-censored, time-to-event data with covariates. Significant differences in pain tolerance between patients with fibromyalgia vs. healthy controls were observed. While the effects of psychological and clinical variables appeared limited, unique contributions of opioid medication use may influence cold pain tolerance in a proportion of patients with fibromyalgia who take opioids.

### 4.1. Reduced CPT survival in fibromyalgia and potential contributors

Confirming previous findings of how individuals with fibromyalgia demonstrate lower tolerance to painful stimuli,^8–10^ in our dataset, individuals with fibromyalgia showed lower rates of survival in the CPT assessment. Under both p-value-based and Lasso regression variable selection methods, fibromyalgia status was the most significant predictor of CPT survival probability even when accounting for psychological measures including anxiety, depression, and negative affect. This suggests that fibromyalgia may impair cold pain tolerance in ways beyond such psychological processes alone, for example, in conjunction with underlying neurobiological mechanisms. In this study, neuropathic measures of chronic pain that may be potentially related to pain tolerance were not evaluated. However, pain sensitivity in fibromyalgia may be attributed to biological alterations spanning both peripheral and central nervous system.^43,44^ Among different patients, fibromyalgia can be characterized by varying amounts of peripheral nociceptor pathologies^45,46^ and central alterations such as altered descending control of pain and central sensitization.^47,48^ The sensation of cold pain, specifically, is mediated by transient receptor potential melastatin 8 (TRPM8), and alterations to this receptor may account for cold hyperalgesia in a variety of chronic pain conditions, including fibromyalgia.^49,50^ Nerve fiber diameter, collected via skin biopsy, has also been shown to be reduced in some fibromyalgia patients.^51^ In future studies of cold pain tolerance, measures of neuropathic pain should be collected in both patients and healthy controls, to complement questionnaire measures.

### 4.2. CPT survival, Behavioral Activation System, and reward response

Overall, the psychological and clinical questionnaire measures collected in the studies were not found to have clear relationships with CPT survival. However, some relationships became more distinct when survival analyses were performed separately for fibromyalgia or healthy control groups. For instance, higher scores in BAS drive and BAS fun-seeking were slightly predictive of higher CPT survival among fibromyalgia patients, but not among healthy individuals.

These results suggest that the psychological contributors to pain tolerance may differ between fibromyalgia patients and healthy individuals. For example, pain tolerance in fibromyalgia patients, compared to healthy controls, may be more susceptible to individuals’ intrinsic drive to achieve goals (i.e., BAS drive), and/or willingness to engage in unknown but possibly rewarding activities (i.e., BAS fun-seeking). Previous research has proposed the BIS-BAS model of chronic pain, suggesting that pain responses can be predicted by individual perceptions of reward and punishment.^52^ Furthermore, prior neuroimaging research in patients with fibromyalgia has demonstrated that BAS drive is related to differences in regional brain response to anticipation of rewards^27^ and avoidance of punishment.^29^ Thus, alterations in recruited CNS activities ongoing during the CPT assessment may contribute to the observed differences in CPT survival between healthy controls and patients with fibromyalgia.

### 4.3. Effects of opioid medication use

CPT survival times were significantly longer in opioid-taking patients compared to non-opioid-taking patients in one cohort of patients (Study 2), but not both. Furthermore, there was no evidence to suggest that these differences could be explained by patients’ daily dose of opioid medication, duration of opioid medication use, or the timing of their last opioid intake. Meanwhile, in line with CPT survival relationships with BAS measures as described above, it is possible that changes in reward response in patients with fibromyalgia who take opioids^29^ may relate to increased cold pain tolerance. Nonetheless, the observed inconsistencies across studies suggest that the relationship between opioid medication use and cold pain tolerance in fibromyalgia patients is tenuous, in that it may vary across patient cohorts and requires further investigation. Prior research has shown that individuals (with and without chronic pain) taking opioids demonstrate lower cold pain tolerance when measured prior to scheduled opioid dose.^53^ In another study, compared to non-opioid patients, opioid-taking patients with chronic pain had similar cold pain tolerance when measured within several hours after their morning analgesic dose.^54^ The timing of opioid dose prior to CPT was variable in the present analysis, therefore, timing effects of opioids are unclear, yet given such prior evidence, opioid timing may be an important variable in assessing cold pain tolerance. Additionally, other studies in individuals with opioid use disorder (i.e., opioid addicted individuals without noted presence of chronic pain) have shown decreased opioid tolerance (by survival analysis) when individuals were tested at a time when opioids (e.g., heroin, methadone) were present in their system.^55^ Together, these prior research findings suggest potential differences in opioid effects on cold pain tolerance in patients with vs. without chronic pain.

### 4.4. Limitations

In this analysis, two separate studies were combined to maximize statistical power. However, differences between the two studies were found to significantly affect survival rates in the CPT. Study 2, in which the testing temperature was standardized to 5° C, was associated with lower CPT hazard compared to Study 1 (with individualized temperatures) when accounting for group status. Although the group effect was the most salient predictor of CPT survival in both studies (when each study was analyzed separately), the study effect in the combined analysis was nearly as notable as the group effect. Such significant differences in CPT survival between studies suggest limitations regarding generalizability of CPT findings across different geographic/demographic cohorts and indicate that slight differences in CPT procedures (i.e., water temperature) may influence generalizability of results across studies comparing fibromyalgia patients. Additionally, it is important to note that the within-group analyses for relationships of clinical/psychological factors (excluding opioid medication use, PANAS negative affect, number of pain areas, and STAI state anxiety) with CPT survival were exploratory, none of which were significant after correcting for multiple comparisons (which may be attributed to a lack of power). Lastly, as described above, timing of opioid intake prior to the CPT assessment in patients who take opioids may be an important contributor to evaluation of cold pain tolerance.

### 4.5. Conclusions

Through using survival analysis to analyze time-to-event CPT data more suitably, the results of this analysis confirm previous findings that, compared to healthy individuals without chronic pain, individuals living with fibromyalgia demonstrate lower tolerance to cold pain. To further understand the relationship between clinical/psychological factors and cold pain tolerance in individuals with fibromyalgia, future investigations should include measures to evaluate the presence of peripheral neuropathy and comparisons of CPT to other tests of CNS and pain processing function in the same patients. Furthermore, the results suggest a potential effect of opioid medication on increasing cold pain tolerance in some fibromyalgia patients. The effect of increased cold pain tolerance in a subset of patients with fibromyalgia who are taking opioids may be related to differences in reward response and motivation and regionally influenced variables such as prescribing practices, and it requires further investigation with timed opioid intake approaches.

## Data Availability

The data that support the findings of this study are available from the corresponding author, K.T.M., upon reasonable request.

## 6. Acknowledgements

We thank Erin Perrine, Christina Cojocaru, and Elizabeth Cha for their assistance with recruitment, data collection, data organization, and analysis for Study 1. We thank Lindsie Boerger, Meghna Nanda, and Drs. Anne Baker and Su Hyoun Park for their assistance with recruitment, data collection, and data organization for Study 2. Special thanks to Dr. Sean Mackey, Stanford University for resources supporting Study 1 data collection. Lastly, we thank all the study participants for their time and contribution to advance clinical research and knowledge.

## 9. Author Contributions

K.T.M. designed the study, analyzed the data, and edited and revised the manuscript. E.Z.D. designed the study, analyzed the data, wrote, edited, and revised the manuscript. D.P.W. provided biostatistical support for study design, data analysis, and editing of the manuscript. All authors have read and approved the final version of the manuscript.

